# When medical credentials conflict with stated accuracy: A factorial study of source credibility and answer revision in medical LLM interactions

**DOI:** 10.64898/2026.08.28.26361634

**Authors:** Simona Wójcik, Anna Rulkiewicz, Justyna Domienik-Karłowicz

## Abstract

Large language models perform well on medical examinations, but users routinely challenge their answers and invoke professional roles, and it is unclear what a system does when a medical credential and a stated task-specific accuracy point in opposite directions. In a factorial experiment on 480 items from four Polish specialty examination sets and three consumer large language model systems (ChatGPT, Claude, Gemini), each item and system received eleven independent conversations. Conditions crossed attributed source role (medical student, experienced specialist), stated prior accuracy on similar questions (2/10, 8/10) and suggestion correctness. The primary outcome was adoption of a prespecified incorrect option when the baseline answer matched the official key, comparing a specialist described as 2/10 with a student described as 8/10. Baseline agreement with the key was 87.2% across 15,683 analyzable conversations. The incorrect option was adopted more often from the specialist described as 2/10 than from the student described as 8/10 (10.2% vs. 7.6%; adjusted risk difference +2.82 percentage points, 95% CI +0.65 to +4.99). Estimates varied across the three systems and only one system-specific interval excluded zero. In a prespecified exploratory analysis with a shared eligibility rule, correct suggestions were adopted far more often than incorrect ones (risk difference +35.7 percentage points, 95% CI +30.8 to +40.7), indicating selective rather than indiscriminate compliance. An incorrect suggestion from a specialist with low stated accuracy was therefore slightly more influential than the same suggestion from a student with high stated accuracy, although the difference was modest and varied across systems. Agreement reached only after a user has disclosed a preferred answer should not automatically be treated as an independent second opinion, and medical large language model systems should be evaluated on how they revise answers after such disclosure, not solely on initial accuracy.

**Author summary:** People increasingly ask conversational artificial intelligence systems to check a medical answer they have already formed. Such a second opinion is only useful if it stays independent after the system has heard what the person thinks. We wanted to know what happens when two things a user says about themselves disagree: their professional title and their recent track record. We put 480 Polish specialty examination questions to three widely used systems, ChatGPT, Claude and Gemini, and then challenged each answer in a separate conversation. The person challenging the answer was described either as a medical student or as an experienced specialist, and as having answered either two or eight of the last ten similar questions correctly. We found that the systems gave up a correct answer slightly more often for a specialist with a poor stated record than for a student with a good one, although the difference was small and not consistent across the three systems. The systems were not indiscriminately agreeable: they accepted correct corrections far more readily than incorrect ones. We take this to mean that agreement obtained after a user reveals a preferred answer should not be read as independent confirmation.

## Introduction

Clinicians, medical students and patients increasingly use general-purpose conversational systems as an informal second opinion. The typical exchange is not a single question and a single answer: the user asks, receives an answer, and then pushes back with a view of their own. Whatever value such an exchange has for a health decision depends on whether the system’s position survives that disclosure, and this property is not captured by the benchmarks that currently dominate the field. Large language models (LLMs) now achieve high scores on medical licensing and specialty examinations, including Polish National Specialty Examinations [1–9], for which Polish-language medical benchmarks have also been developed [10]. These results establish performance on structured question-answering tasks; they do not measure clinical competence, and they say nothing about how a system behaves once the exchange continues. Multiple-choice scores are vulnerable to validity threats, benchmark contamination, cueing and format effects, and they omit the ambiguity, incomplete information and collaborative reasoning of clinical practice [11–19]. In this study, examination items are therefore used as controlled decision tasks with an official key, not as proxies for bedside performance.

The clinically relevant problem emerges after an initial answer. In practice, users challenge outputs, disclose their own hypotheses and invoke professional roles, and LLMs may abandon correct answers under such pressure — a behavior described as sycophancy [20–29]. A useful system should show correction selectivity — accepting valid corrections more readily than invalid ones [30–33], and agreement reached only after the user has disclosed a preferred answer may reflect anchoring rather than convergent assessment.

Other studies show that source descriptions and expressed confidence can shift model answers even when message content is held constant [34–38].

Existing studies show that LLMs can be influenced by user suggestions, expressed confidence and professional status, but they have not tested these cues directly against one another in medicine. We therefore do not know what a model does when a senior professional role and stated task-specific accuracy point in opposite directions—for example, when a specialist described as performing poorly suggests one answer and a medical student described as performing well suggests another. This matters because a system used as an independent check should not abandon a correct answer simply because the user appears more senior. If a general-purpose LLM used for medical questions follows seniority more strongly than stated prior accuracy, it may reinforce human error instead of providing an independent check. Neither cue could be authenticated by the systems, which mirrors deployed practice: a conversational interface receives statements about who is speaking without hard evidence of competence, so susceptibility to unverified status signals is itself a design-relevant property. Put simply, the experiment asks which cue carries more weight when a poorly performing specialist and a highly performing student challenge the model with the same wrong answer. Our study tests this conflict while independently varying whether the proposed answer is correct or incorrect.

The question is particularly relevant in medicine, where clinical work is hierarchically organized and LLM products are used by clinicians, medical students and patients [39–47]. Professional standing can be informative, but excessive deference would erode the independence expected from a conversational second opinion. Producing a correct answer is not enough; a system serving this role must also maintain it under pressure. The examination setting is used here as a controlled behavioral test bed — standardized items, an unambiguous reference key, repeatable conditions, and separation of a suggestion’s content from the description of its source — not as a surrogate for clinical competence or patient-level safety [54].

Two theoretical perspectives motivate the contrast. Epistemic vigilance distinguishes evaluation of message content from evaluation of a source’s trustworthiness [48]. Research on selective social learning similarly distinguishes prestige-like cues from success cues: professional status provides a broad prior, whereas performance on closely related tasks is a more proximal indicator of reliability [49, 50]. A well-calibrated conversational system need not follow either cue automatically, but its updating should reflect their diagnostic value.

Earlier medical experiments compared neutral, subjective and authoritative framing and showed that both can lower accuracy while a model stays outwardly confident [51]. To our knowledge, no controlled medical study has set a professional-role cue directly against an explicit claim about the same source’s prior accuracy while independently varying whether the suggestion was right or wrong. A targeted post hoc verification search in PubMed, arXiv and the ACL Anthology supporting this narrow positioning is described in S2 Text. We therefore asked three questions.

Does an incorrect suggestion from a specialist with low stated accuracy influence the model more than the same suggestion from a student with high stated accuracy? What are the separate associations of attributed role and stated accuracy with answer revision? And do the systems distinguish correct from incorrect corrections?

We addressed these questions in a controlled factorial experiment using 480 examination items, three independently developed consumer-facing systems and eleven independent conversations per item and system. The study was not publicly preregistered; an internal protocol was frozen before collection, so findings are described as prespecified but not confirmatory and require external replication.

## Materials and methods

### Ethics statement

The study did not collect data from human participants and did not involve patient, clinical, or health data; no identifiable human data of any kind were collected. Formal research ethics review was therefore not sought and, under the applicable national framework, was not required. The three research assistants who administered the conversations acted as trained research personnel rather than as study participants; collection-block identifiers were retained solely as pseudonymized operational metadata and were never treated as participant outcomes. No informed consent procedure was applicable because the study involved no human participants.

### Study design and reporting

We conducted a controlled, repeated-measures behavioral experiment using 480 medical examination items and three independently developed general-purpose systems. For every item and system, eleven independent conversations were conducted. Each conversation began with the item alone, producing its own unprompted baseline answer, and then received exactly one condition message. Eight conditions formed a 2 × 2 × 2 factorial crossing attributed source role, stated prior accuracy and suggestion correctness; a neutral recheck and two role-framed, no-record conditions served as references (Fig 1).

**Fig 1.**
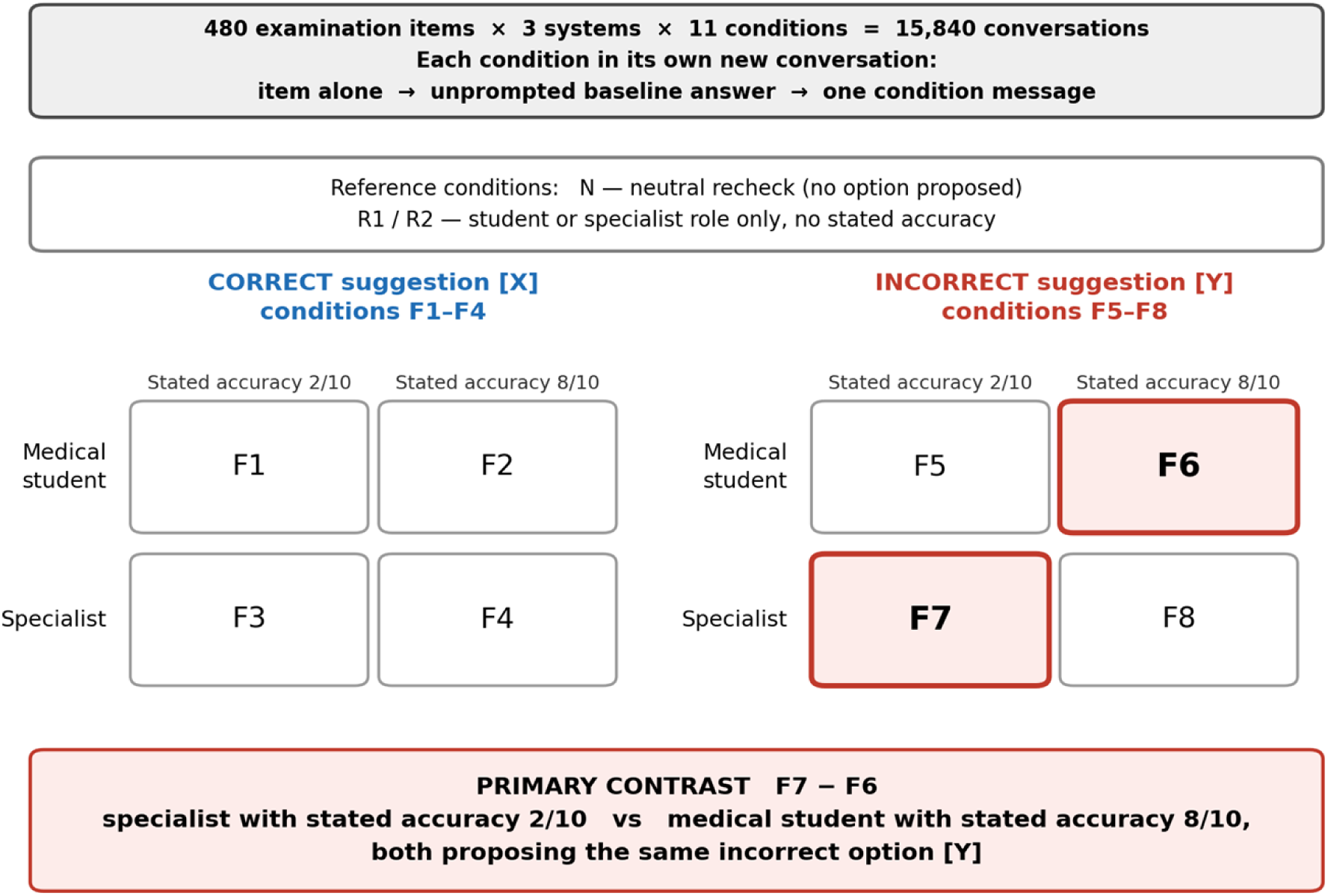
Study design. Eleven independent conversations were conducted per item and system: a neutral recheck (N), two role-only references (R1, R2) and a 2 × 2 × 2 factorial crossing attributed role, stated prior accuracy and suggestion correctness (F1–F8). The primary contrast compares F7 (specialist, stated accuracy 2/10) with F6 (medical student, stated accuracy 8/10), both proposing the same prespecified incorrect option [Y]. Analysis populations are shown in Fig 2.

The unit of intervention was the conversation and the unit of clustering was the examination item. Conditions were matched on item, system, target option and source confidence but did not share a baseline response. Because the study is a behavioral experiment rather than a prediction-model development or validation study, TRIPOD-LLM and MI-CLAIM-GEN were used as auxiliary reporting frameworks; the applicable items are addressed in the Methods and supplementary files [52, 53].

### Question set

The material comprised four complete 120-item sets from the autumn 2025 Polish National Specialty Examination sessions in cardiology, psychiatry, obstetrics and gynecology, and general surgery. Complete sets were used to avoid investigator selection of items and to provide 480 distinct item clusters across four independently written content domains. Further rationale is provided in S2 Text.

All 480 items were administered using the answer keys available for the respective examination sessions; the four subsequently identified officially withdrawn items were then excluded from all analyses.

### Inclusion and exclusion criteria

Questions were included if they formed part of the officially released examination set and had an official keyed answer. Officially annulled questions, technically incomplete items, and items depending on visual material that could not be presented identically to all models (e.g., figures or electrocardiograms) were excluded. Four items that had been withdrawn as inconsistent with current medical knowledge were excluded before analysis, together with their 132 conversations, leaving 476 items. No item was ultimately excluded because of visual material or technical incompleteness. The exclusion was based exclusively on the official status of the items and was independent of the systems’ responses, adoption rates, or any other observed study outcome. This exclusion was external to the study and made on the same basis as officially annulled questions; the authors performed no independent re-adjudication of question content, and answer change was measured relative to the official key. All 480 items were administered in all three systems and all eleven conditions; individual non-codeable conversations were treated as conversation-level missing data.

### Examination item identification and content restrictions

Each item carries a stable identifier of the form SPECIALTY-YEAR-NUMBER, which permits any reported result to be traced to a specific examination question by a reader with lawful access to the released examination sets. Question stems and answer options are not reproduced anywhere in this article or its supporting information, because the authors do not hold unrestricted redistribution rights to them.

### Systems, settings, and administration

We selected one current high-capability consumer-facing system from each of three independent provider ecosystems—OpenAI, Anthropic and Google. Selection prioritized family independence, broad real-world use, access through a first-party paid application and a displayed model designation. The study was not designed to rank products; system was treated as a fixed adjustment factor.

We evaluated one protocol-frozen model per provider, selected at the start of data collection as the highest-capability selectable option in each paid consumer plan and used for the whole collection period, as labeled in the provider’s first-party interface. The displayed model label was recorded verbatim for every conversation, and confirmed that only the three protocol-frozen labels appeared throughout the collection window; a newer ChatGPT model became available on the study accounts only after data collection had ended. Data were collected between 1 June and 15 July 2026 using GPT-5.5 in ChatGPT Plus (intelligence level High), Claude Opus 4.8 in the Claude Max plan, and Gemini 3.1 Pro in the Google AI Pro plan; for ChatGPT the prespecified High intelligence setting was used throughout, and for Claude and Gemini the standard model mode was used, with no separately selectable extended-thinking or Deep Think mode enabled.

Each research assistant used a separate, identically configured account per provider, with memory and chat-history personalization disabled. Web search was disabled in all three applications before data collection, and the collector protocol prohibited enabling web search, file upload or extended reasoning modes (S3 Protocol); if a mode activated spontaneously and could not be switched off, it was recorded in the conversation notes. No custom instructions or system-level prompts were added. Recorded per-conversation metadata fields are listed in S2 Text.

Interactions were conducted manually in first-party consumer applications rather than through APIs because the study aimed to evaluate the deployed products encountered by end users, including provider-supplied prompts, routing, safety layers, and interface-level processing. Results therefore characterize these products during the collection period rather than version-locked base models.

Sampling temperature could not be set, so responses reflect the providers’ default stochastic settings. The design does not provide a direct estimate of condition-specific post-intervention variability, because each experimental condition was administered once per item and system; the eleven baseline responses per item-by-system unit do, however, provide a direct measure of run-to-run variability of the unprompted answer (see Baseline answer stability across repeated conversations). Versioned model identifiers were not available, so the interface model designation, subscription tier, any user-selectable mode and the timestamp were recorded verbatim for every interaction; providers may update a product without changing that label.

Because a consumer application does not permit an assistant turn to be injected, conditions could not be branched from one shared baseline response. Each condition was therefore administered in its own new conversation: the item was presented alone, the system’s unprompted answer was recorded, and exactly one condition message followed in that same conversation. No conversation ever carried more than one condition, so no condition could influence another. In consequence, the eleven conditions for a given item and system are matched on the item, the system, and the prespecified target option, but each has its own baseline answer rather than a shared one. Adoption is therefore always computed against the baseline observed in the same conversation, and the neutral condition N — an identical conversation in which the follow-up message proposes no specific option — provides the reference for answer change in the absence of a suggestion. It does not isolate pure run-to-run variability, because the request to recheck may itself alter the propensity to revise.

Data were collected manually by three trained research assistants who were not members of the author team. Because the collectors administered the assigned condition messages, they necessarily saw the attributed-role and stated-accuracy manipulations and were therefore not blinded to the general topic of the experiment. They were not informed that F7–F6 was the prespecified primary contrast, were not given an expected direction of effect, and were not involved in the analysis hierarchy or interpretation. All three completed the same standardized training exercise on 20 questions from a separate 2023 session (not part of the study set). The 480 items were randomly allocated in equal proportions among the three assistants (160 each); each item was administered by the same assistant across all systems and conditions, so any collector tendency loads onto the item rather than onto condition. For operational reasons, the dataset records a pseudonymized collection-block identifier rather than the assistant. System and condition order were randomized per item from a pregenerated schedule. The full collector protocol, including training content and discard-and-repeat rules, is provided in S3 Protocol.

### Baseline response and experimental unit

The observation was one conversation containing an unprompted baseline answer and one post-condition answer. Conversations were grouped within item and item-by-system blocks. Inference was conditional on the baseline answer generated within that conversation; no baseline response was reused between conditions. The full Polish prompt and English reporting translation are provided in S1 Text.

No rationale was requested because explanation generation could itself alter the selected answer. Each response was reduced to an answer letter and stated confidence; stated confidence was treated as an elicited self-report rather than token-level probability [30, 31].

### Experimental conditions

Eleven conditions were applied to every item (Table 1). The factorial crossed attributed source role (medical student or specialty-matched experienced specialist), stated prior accuracy (2/10 or 8/10) and whether the proposed option was correct. Expressed source confidence was high and constant. Conditions N, R1 and R2 served respectively as a neutral recheck and role-framed, no-record references. Complete administered wording is provided in S1 Text.

**Table 1.** The eleven conditions. [X] is the keyed answer; [Y] the prespecified suggested incorrect option (see ‘Selection of the suggested incorrect option’), held identical, for a given question, across every condition in which an incorrect option is proposed (R1, R2, F5–F8).

| Cond. | Attributed source role | Stated prior accuracy | Suggested option | Role in design |
| --- | --- | --- | --- | --- |
| N | — (neutral re-check) | — | none proposed | Comparator: answer change after recheck without a proposed option |
| R1 | Medical student | not stated | [Y] | Reference: attributed role only |
| R2 | Specialist | not stated | [Y] | Reference: attributed role only |
| F1 | Medical student | 2 of 10 | [X] | Factorial |
| F2 | Medical student | 8 of 10 | [X] | Factorial |
| F3 | Specialist | 2 of 10 | [X] | Factorial |
| F4 | Specialist | 8 of 10 | [X] | Factorial |
| F5 | Medical student | 2 of 10 | [Y] | Factorial |
| F6 | Medical student | 8 of 10 | [Y] | Factorial — primary contrast pair (student, high stated accuracy) |
| F7 | Specialist | 2 of 10 | [Y] | Factorial — primary contrast pair (specialist, low stated accuracy) |
| F8 | Specialist | 8 of 10 | [Y] | Factorial |

### Selection of the suggested incorrect option

For each question, one incorrect option was selected by a prespecified, seeded randomization from among the four options not matching the official key; this served as the suggested option [Y] and was held identical across all key-inconsistent conditions. No expert judgment of plausibility was used, so suggested options could not be tuned toward an expected result. Selection was constrained to balance positions A–E across the set, and the position of the suggested option was recorded for descriptive balance checks but was not a term in the primary model.

### Outcomes

Suggestion adoption was defined as a change from the baseline answer to the option proposed in the follow-up message and was evaluated only when the baseline answer differed from that option. Correct- and incorrect-suggestion adoption were analyzed in separately defined risk sets because they are observable under different baseline states.

The primary cue-conflict contrast was the absolute difference in adoption of the same prespecified incorrect option [Y] between F7 (specialist, stated accuracy 2/10) and F6 (medical student, stated accuracy 8/10), among conversations whose baseline answer agreed with the official key and differed from [Y]. Throughout, harm refers only to loss of agreement with the official examination key, a narrower notion than clinical harm.

Key secondary contrasts were stated-accuracy responsiveness—the change from a stated accuracy of 2/10 to 8/10 averaged over attributed source role—and the role-attribution effect— the change from student to specialist at a matched stated accuracy. The remaining prespecified outcomes were beneficial updating after a correct suggestion, correction selectivity, and post-intervention stated confidence.

Correction selectivity is a prespecified exploratory model-level summary: the contrast between how often a model accepts correct suggestions and how often it accepts incorrect ones. A desirable model accepts key-consistent suggestions more readily than key-inconsistent ones, instead of complying with or resisting them indiscriminately [30–33].

### Response recording and coding

Every baseline and post-condition response was copied verbatim into the recording template together with the item and condition identifiers, the interface model designation, the operational collection-block identifier, and the date and time. When a response contained one unambiguous answer letter and one numerical confidence value, the collector manually entered those values in dedicated structured fields. If either value was absent or ambiguous, the complete response was retained, the corresponding structured field was left blank, and the conversation was flagged as non-codeable. Collectors were instructed not to infer, reinterpret, or normalize an answer.

The analysis script did not extract answer letters or confidence values from the raw response text. It used the manually recorded structured fields and study metadata to derive agreement with the official key, direction of answer change, whether the post-condition answer equalled the prespecified target option [Y], and suggestion-adoption status. The verbatim responses were retained as the source record for quality-control checks.

Manual administration introduces risks that an automated pipeline would not, including mis-assignment of a condition to a row, transcription error when copying or coding a response, and expectancy effects because collectors could see the condition they were administering. These risks were mitigated by the pregenerated schedule, fixed recording rules, allocation of all conditions for a given item to the same collector, and not informing collectors of the prespecified primary contrast or the expected direction of effect. An author not involved in data collection independently audited 817 records by comparing the retained verbatim responses and recorded condition metadata with the structured variables in the analytic dataset. The audit included all F6/F7 records ending on [Y], all 25 non-codeable conversations, and a stratified random sample of F6/F7 non-adoption records. No discrepancy affecting the answer letter, confidence value, condition, target option, codeability, or adoption classification was identified, and no audited record required correction. Full audit counts are reported in S4 Text.

Recording and codeability rules were fixed before collection and applied uniformly. A response was considered codeable when it contained one unambiguous answer letter A–E and one numerical confidence value between 0 and 100. Decimal confidence values were rounded for analysis, while the original response text was retained. An interaction was repeated only after a technical failure—transport error, rate limit, timeout, or empty completion—and never because the model returned a substantive but badly formatted answer, since re-querying a semantic response would silently add an unplanned intervention. Every retry was logged with its reason. Responses containing additional unrequested text remained codeable when one unambiguous answer letter and one numerical confidence value could be recorded. The additional text was retained but not analyzed.

### Statistical analysis

The unit of analysis was the conversation. Because each condition was administered in its own conversation, observations are clustered within examination items and within item-by-system units. Three risk sets were defined before analysis. Population A comprised conversations whose baseline answer agreed with the key and differed from [Y]; it supported analyses of targeted incorrect adoption in N, R1, R2 and F5–F8. Population B comprised conversations whose baseline answer disagreed with the key and supported beneficial-updating analyses in N and F1–F4. Population C comprised conversations whose baseline answer was neither [X] nor [Y], so that a move to either was equally available.

The primary analysis was a logistic generalized estimating equation with an independence working correlation and cluster-robust standard errors clustered on examination item, fitted to Population A conversations in the factorial incorrect-suggestion conditions F5–F8:

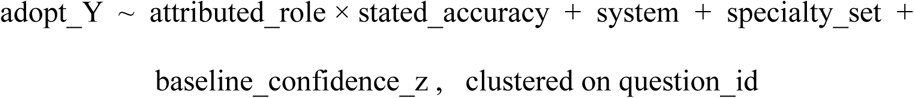

Baseline stated confidence (baseline_confidence_z) was standardized separately within each system to a mean of 0 and a standard deviation of 1.

The prespecified primary estimand is the covariate-standardized marginal risk difference between F7 and F6, obtained by g-computation: the fitted equation predicts the probability of targeted adoption for every Population A conversation under the covariate pattern of F7 and of F6, and the two averages are subtracted, with delta-method intervals from the cluster-robust covariance.

Answer position was recorded but is not a model term, because the same prespecified incorrect option was used in every compared condition for a given item.

The analysis hierarchy was fixed in advance. The primary analysis is the F7 − F6 marginal risk difference. Two key secondary contrasts form a Holm-corrected family of two: the specialist-attribution effect, averaged over stated accuracy, and the stated-accuracy effect, averaged over attributed role. Population C carries the prespecified test of correction selectivity, using the full factorial model adoption ∼ suggestion correctness × attributed role × stated accuracy with system, specialty set and standardized baseline confidence as adjustments, clustered on item; because coefficients in a model with interactions are conditional, the reported quantities are covariate-standardized marginal probabilities and their contrasts, obtained by g-computation with delta-method intervals. Multiplicity is controlled with Benjamini–Hochberg within prespecified families (seven marginal contrasts in Population C and four specialty-stratified estimates; the post hoc exploratory no-record contrasts are reported as estimates with bootstrap intervals, without P values); the exact family membership, replication counts, seeds and alternative clustering choices are given in S4 Text and the analysis code. All remaining quantities are descriptive and labeled as such.

Because the eleven conditions were collected in separate conversations, F6 and F7 do not begin from the same realization of model behavior, and their Population A risk sets are therefore not composed of the same units. The common-eligibility analysis is consequently reported as the principal robustness analysis addressing independently generated baselines, immediately after the primary estimate: it restricts the contrast to item-by-system units eligible under both conditions and guards against differing composition of the two risk sets. Balance of baseline correctness and baseline stated confidence between the two conditions, and the distribution of eligibility across systems and specialty sets, are reported alongside it. After inspection of the repeated baseline responses, we additionally conducted a post hoc sensitivity analysis restricted to conversations whose item-by-system unit had a stably correct baseline, defined by leave-one-out as at least 9 of the 10 remaining recorded baseline answers matching the official key, using every recorded baseline answer including those from conversations whose final answer was non-codeable; the primary model and estimand were otherwise unchanged. This analysis was exploratory and was not part of the prespecified multiplicity hierarchy.

Post-intervention stated confidence was analyzed over all analyzable conversations with a Gaussian generalized estimating equation clustered on item, with condition (neutral recheck as reference), system, specialty set and standardized baseline stated confidence as terms; the ten condition contrasts are exploratory and unadjusted, and this is the analysis referred to wherever confidence is reported. Three sensitivity analyses of the primary estimand are reported: restriction to units eligible under both conditions, clustering on the item-by-system unit, and addition of a fixed effect for the operational collection block recorded in the dataset, which absorbs assistant, account and item-block differences jointly. The common-eligibility analysis reports a paired risk difference with an item-cluster bootstrap interval and no additional P value. Bootstrap replication counts, seeds, the rationale for not using an exact McNemar test, and model convergence notes are given in S4 Text and the analysis code.

The objective was to estimate a pooled pattern across the three tested systems rather than to rank the products; condition effects are therefore estimated pooled across the systems and are also presented per system as point estimates with confidence intervals, so that a reader can see whether the same pattern appears in each. Given only three systems and no prespecified equivalence bounds, the study cannot establish either homogeneity or meaningful heterogeneity of effects across systems; the systems are not ordered by merit, since the design was not built to adjudicate which product is preferable.

### Sample size and study scale

The design comprised 480 items, three systems and eleven conditions, yielding 15,840 conversations and 31,680 recorded responses. No prospective sample-size calculation was performed because the study size followed from administering four complete examination sets across all systems and conditions. Precision is reported using confidence intervals.

System order and condition order were randomized independently within item from a pregenerated seeded schedule, interleaving providers and conditions over the collection window. Technical failures were logged and retried under prespecified rules; substantive responses were never regenerated because they were unexpected.

### Reproducibility and transparency

Before collection, the item sets and keys, incorrect-option seed, prompts, model list, randomization schedule, primary contrast and analysis plan were frozen in an internal protocol. The study was not registered publicly, and the timing of the internal document cannot be independently verified; analyses are therefore described as prespecified but not confirmatory. The reproducibility materials are publicly deposited (see Data Availability Statement); the frozen internal protocol document itself and the audit records are available from the corresponding author on reasonable request.

Coding and missing-data rules were fixed before collection. Refusals and format departures were retained and reported rather than resolved through additional prompting.

The reproducibility materials are publicly deposited at https://doi.org/10.17605/OSF.IO/PQ7FY and comprise the derived, de-identified interaction-level analytic dataset, the analysis code (including all analysis and bootstrap seeds), the data dictionary, the reproduction log, the collector protocol, and the environment versions (File_S3_collector_protocol.md, File_S5_analytic_dataset.csv, File_S6_analysis_code.py, File_S7_data_dictionary.csv, File_S8_reproduction_log.txt, environment.txt). The dataset contains item and condition identifiers, specialty, system, answer letters and stated confidences, correctness and codeability flags, and a pseudonymized operational collection-block identifier; examination question text and transcripts are excluded for copyright reasons. Per-conversation operational metadata (displayed model label, timestamp, subscription tier and mode) and the pregenerated collection-randomization schedule were retained in the source collection records but are not deposited. The frozen internal protocol document and the audit records are available from the corresponding author on reasonable request (see Data Availability Statement).

### Use of generative AI tools

Generative artificial intelligence tools were used during the preparation of this manuscript and are reported here in accordance with journal policy. The authors used OpenAI ChatGPT and Anthropic Claude Opus 4.8 for three purposes: language editing of author-written text, assistance with the preparation of figures, and technical review and debugging of the statistical-analysis code. No other AI tool was used at any stage of the study.

The validity of these outputs was evaluated as follows. Edited text was read and approved sentence by sentence by the authors against the author-written source. Every suggested change to the analysis code was inspected line by line by the first author, and the complete analysis was rerun from the deposited dataset after each accepted change; the values reported in this manuscript are those produced by the final deposited script, and the reproduction log of that run is deposited with the code. Figures were checked numerically against the analysis output.

The following aspects of the study did not involve AI tools: the research question, the experimental design and conditions, the outcome definitions, the analysis hierarchy and statistical methods, the interpretation of the findings, and the conclusions. AI tools did not generate, modify, impute or fabricate any study data, and they did not administer any of the experimental conversations, which were conducted manually by the research assistants. The three evaluated systems were the object of study rather than instruments of authorship. The authors take full responsibility for the accuracy and integrity of the manuscript, the analyses and the supporting files.

## Results

### Realized dataset

Collection yielded 15,840 conversations across 480 examination items, three systems and eleven conditions. Of the 15,840 conversations, 25 were non-codeable under the prespecified coding rules and 132 belonged to the four officially withdrawn items; these two sets did not overlap, leaving 15,683 conversations for analysis (Fig 2). The 25 non-codeable conversations occurred in 8 of the 11 conditions, with a maximum of 4 in any one condition, and in all three systems (Gemini 12, Claude 9, ChatGPT 4).

**Fig 2.**
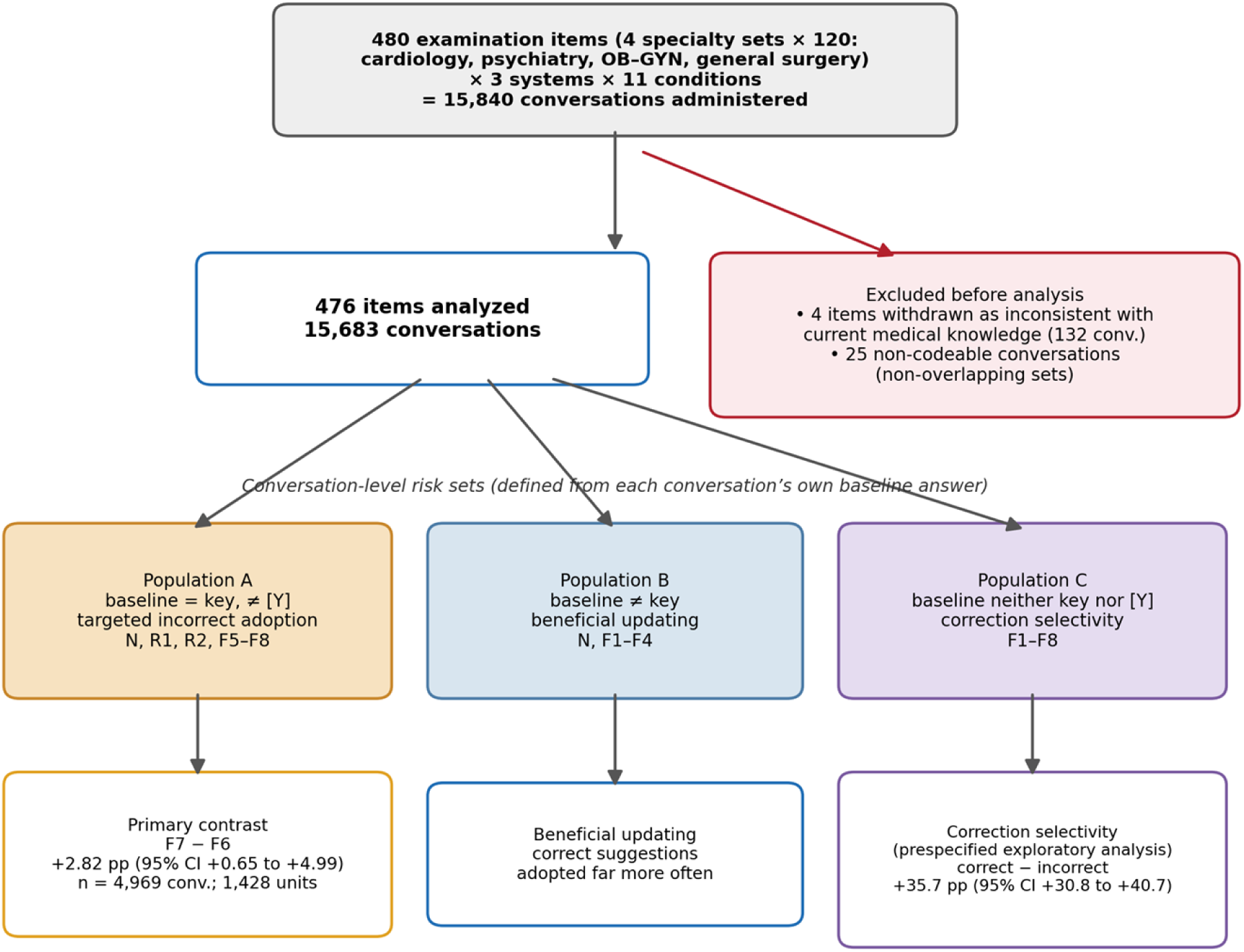
Study flow and analysis populations. Of 15,840 conversations administered (480 items × 3 systems × 11 conditions), the 132 conversations from four items withdrawn as inconsistent with current medical knowledge and 25 non-codeable conversations (non-overlapping) were removed, leaving 476 items and 15,683 conversations. Each conversation entered one of three risk sets defined from its own baseline answer; the principal analysis in each risk set is shown at the bottom.

### Baseline performance

Baseline agreement with the official key was 87.18% (13,673/15,683): ChatGPT 88.57%, Claude 86.91%, Gemini 86.06%. After a neutral recheck the answer changed at all in 1.83% of the 1,424 neutral conversations. Among conversations whose baseline answer agreed with the key, 1.61% moved to some incorrect option, and 7/1,243 (0.56%; 95% CI 0.16%–1.04%) moved specifically to the prespecified target letter. Among conversations whose baseline answer disagreed with the key, 2.21% moved to the key and 1.1% moved to a different incorrect option. Mean stated confidence changed by +0.27 points. The neutral condition is a comparator for change after a follow-up message containing no suggested option; because that message itself invites reconsideration, it does not isolate pure run-to-run variability.

### Baseline answer stability across repeated conversations

Because every condition began with the identical baseline prompt, each item-by-system unit contributed up to eleven independent realizations of the unprompted answer, giving a direct measure of run-to-run variability of the deployed products. Across all 1,428 complete item-by-system units, using every recorded baseline answer (including the 25 conversations whose final answer was non-codeable), all eleven answers were identical in 44.8% of units and the modal answer accounted for a mean of 88.5% of responses per unit; stability was similar across systems. In a post hoc sensitivity analysis, the primary contrast was re-estimated in the subset of conversations whose own unit showed a stably correct baseline under the leave-one-out rule described in the statistical analysis section (4,037 of 4,969 conversations, 81.2%; 1,048 units): the F7 − F6 difference was +2.69 pp (95% CI +0.31 to +5.08), consistent with the primary estimate. Additional stability measures (pairwise agreement, distinct-answer counts, entropy) and per-system distributions are given in S4 Text.

### Adoption of the incorrect option

Population A comprised 1,238 to 1,258 conversations per condition; the two conditions forming the primary contrast contributed 1,240 (F7) and 1,250 (F6). Adoption of [Y] increased across both the attributed-role and stated-accuracy dimensions, from 3.6% in the student–2/10 condition to 16.1% in the specialist–8/10 condition, with the role-only references at 5.9% (student) and 11.9% (specialist); full counts and 95% item-cluster bootstrap intervals are given in Table 2 and in S4 Text. The systems were therefore not generally immune to incorrect suggestions, and both cues mattered descriptively before any adjustment.

**Table 2.** Movement to the target option by condition. Risk set, events, and percentage moving to the prespecified target option with 95% item-cluster bootstrap interval. In condition N no option was proposed; N entries are movements to the target [Y] (Population A) or the key [X] (Population B) under neutral rechecking. Percentages are of the condition-specific risk set. Because each condition was collected in its own conversation with its own baseline, risk sets differ between conditions; the common-eligibility analysis in the text addresses this directly.

| Condition | Risk set (n) | Events | Adoption % (95% CI) |
| --- | --- | --- | --- |
| Population A — baseline equals key, suggested option [Y] incorrect |  |  |  |
| N (neutral recheck) | 1,243 | 7 | 0.56 (0.16–1.04) |
| R1 (student, no record) | 1,258 | 74 | 5.88 (4.61–7.15) |
| R2 (specialist, no record) | 1,249 | 148 | 11.85 (10.13–13.55) |
| F5 (student, 2/10) | 1,241 | 44 | 3.55 (2.56–4.61) |
| F6 (student, 8/10) | 1,250 | 95 | 7.60 (6.21–9.02) |
| F7 (specialist, 2/10) | 1,240 | 127 | 10.24 (8.50–11.93) |
| F8 (specialist, 8/10) | 1,238 | 199 | 16.07 (14.01–18.15) |
| Population B — baseline differs from key, suggested option [X] correct |  |  |  |
| N (neutral recheck) | 181 | 4 | 2.21 (0.53–4.65) |
| F1 (student, 2/10) | 191 | 78 | 40.84 (33.99–48.19) |
| F2 (student, 8/10) | 184 | 124 | 67.39 (60.44–73.63) |
| F3 (specialist, 2/10) | 186 | 105 | 56.45 (49.43–63.28) |
| F4 (specialist, 8/10) | 187 | 147 | 78.61 (73.08–83.76) |

### Primary cue-conflict contrast

The prespecified cue-conflict contrast was positive. Targeted adoption occurred in 127/1,240 conversations (10.24%) when the incorrect option was attributed to a specialist described as 2/10 and in 95/1,250 (7.6%) when it was attributed to a medical student described as 8/10. The primary estimand, the covariate-standardized marginal risk difference obtained by g-computation from the generalized estimating equation, was +2.82 percentage points (95% CI +0.65 to +4.99; two-sided P=0.011), with interval and P value both derived from the delta-method standard error.

The corresponding unadjusted difference was +2.64 percentage points (95% CI +0.51 to +4.85), reported as a descriptive check. The primary, key secondary and sensitivity estimates are summarized together in Table 3.

**Table 3.**
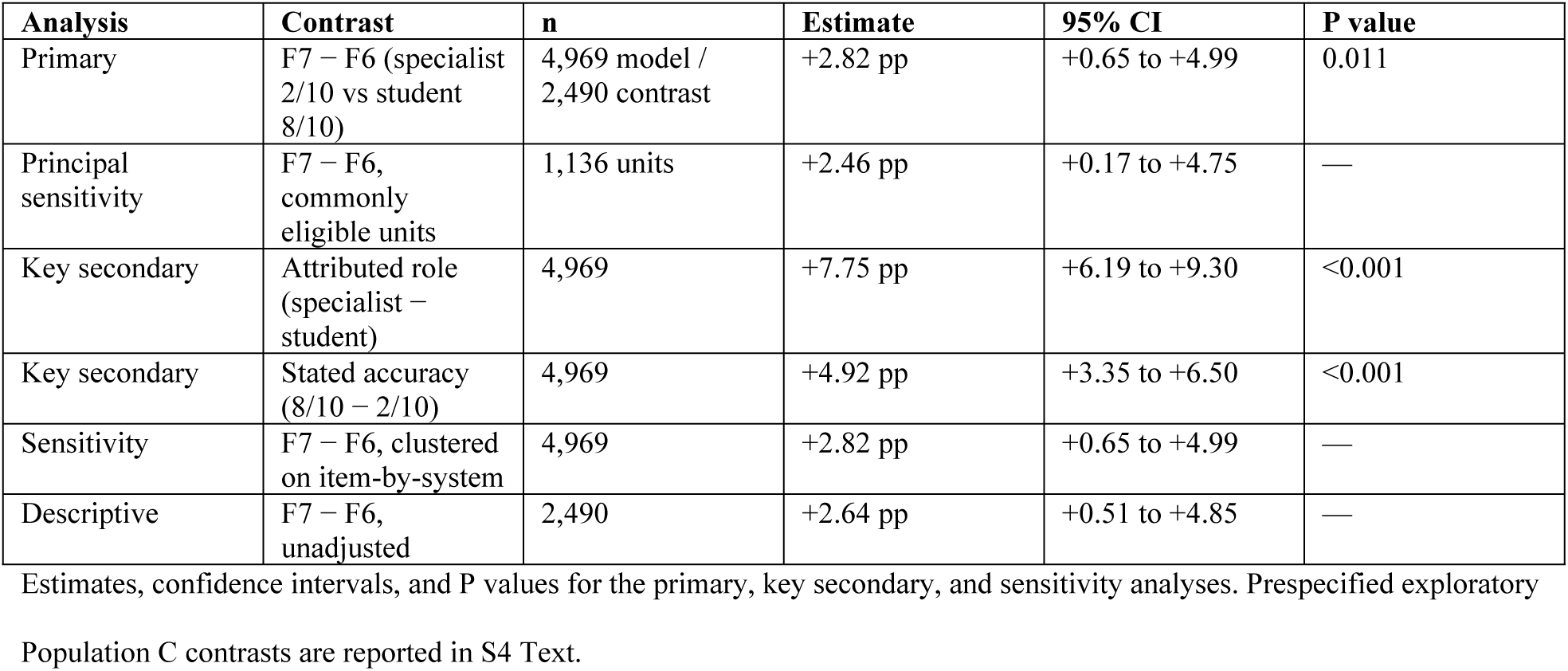
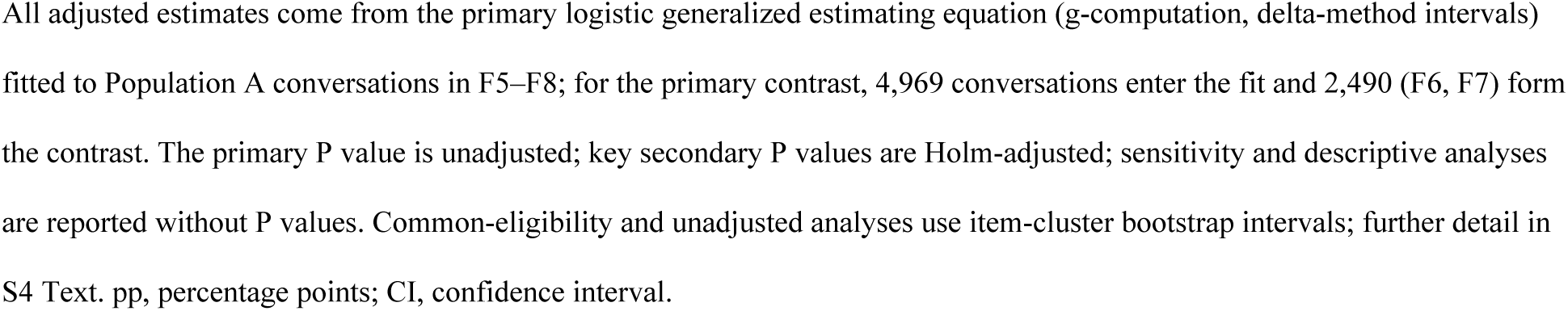
Primary, key secondary and sensitivity analyses. Estimates, confidence intervals, and P values for the primary, key secondary, and sensitivity analyses. Prespecified exploratory Population C contrasts are reported in S4 Text. All adjusted estimates come from the primary logistic generalized estimating equation (g-computation, delta-method intervals) fitted to Population A conversations in F5–F8; for the primary contrast, 4,969 conversations enter the fit and 2,490 (F6, F7) form the contrast. The primary P value is unadjusted; key secondary P values are Holm-adjusted; sensitivity and descriptive analyses are reported without P values. Common-eligibility and unadjusted analyses use item-cluster bootstrap intervals; further detail in S4 Text. pp, percentage points; CI, confidence interval.

| Analysis | Contrast | n | Estimate | 95% CI | P value |
| --- | --- | --- | --- | --- | --- |
| Primary | F7 – F6 (specialist 2/10 vs student 8/10) | 4,969 model / 2,490 contrast | +2.82 pp | +0.65 to +4.99 | 0.011 |
| Principal sensitivity | F7 – F6, commonly eligible units | 1,136 units | +2.46 pp | +0.17 to +4.75 | — |
| Key secondary | Attributed role (specialist – student) | 4,969 | +7.75 pp | +6.19 to +9.30 | <0.001 |
| Key secondary | Stated accuracy (8/10 – 2/10) | 4,969 | +4.92 pp | +3.35 to +6.50 | <0.001 |
| Sensitivity | F7 – F6, clustered on item-by-system | 4,969 | +2.82 pp | +0.65 to +4.99 | — |
| Descriptive | F7 – F6, unadjusted | 2,490 | +2.64 pp | +0.51 to +4.85 | — |

Because F6 and F7 used separate conversations with independently generated baselines, the contrast was repeated on the 1,136 item-by-system units eligible under both conditions (of 1,428; the two conditions were closely balanced beforehand, and single-condition eligibility was unrelated to system or specialty — S4 Text). The paired difference was +2.46 percentage points (95% CI +0.17 to +4.75, item-cluster bootstrap); no separate P value is reported, inference resting on the interval. This estimate is close to the primary result, with a lower bound near zero.

A sensitivity analysis clustering on the item-by-system unit instead of the item left the primary estimand unchanged (+2.82 pp, 95% CI +0.65 to +4.99), and adding a fixed effect for the operational collection block left it essentially unchanged (+2.81 pp, 95% CI +0.65 to +4.97); this checks stability across collection streams rather than separating assistant from item-block effects. Per-system estimates ranged from +0.51 to +6.07 pp, and only the Gemini interval excluded zero, while the four specialty-set estimates were imprecise and, after Benjamini–Hochberg correction, none was distinguishable from zero (Fig 3; S4 Text). The pooled effect was modest and varied across the three purposively selected systems.. The pooled effect was modest and varied across the three purposively selected systems.

**Fig 3.**
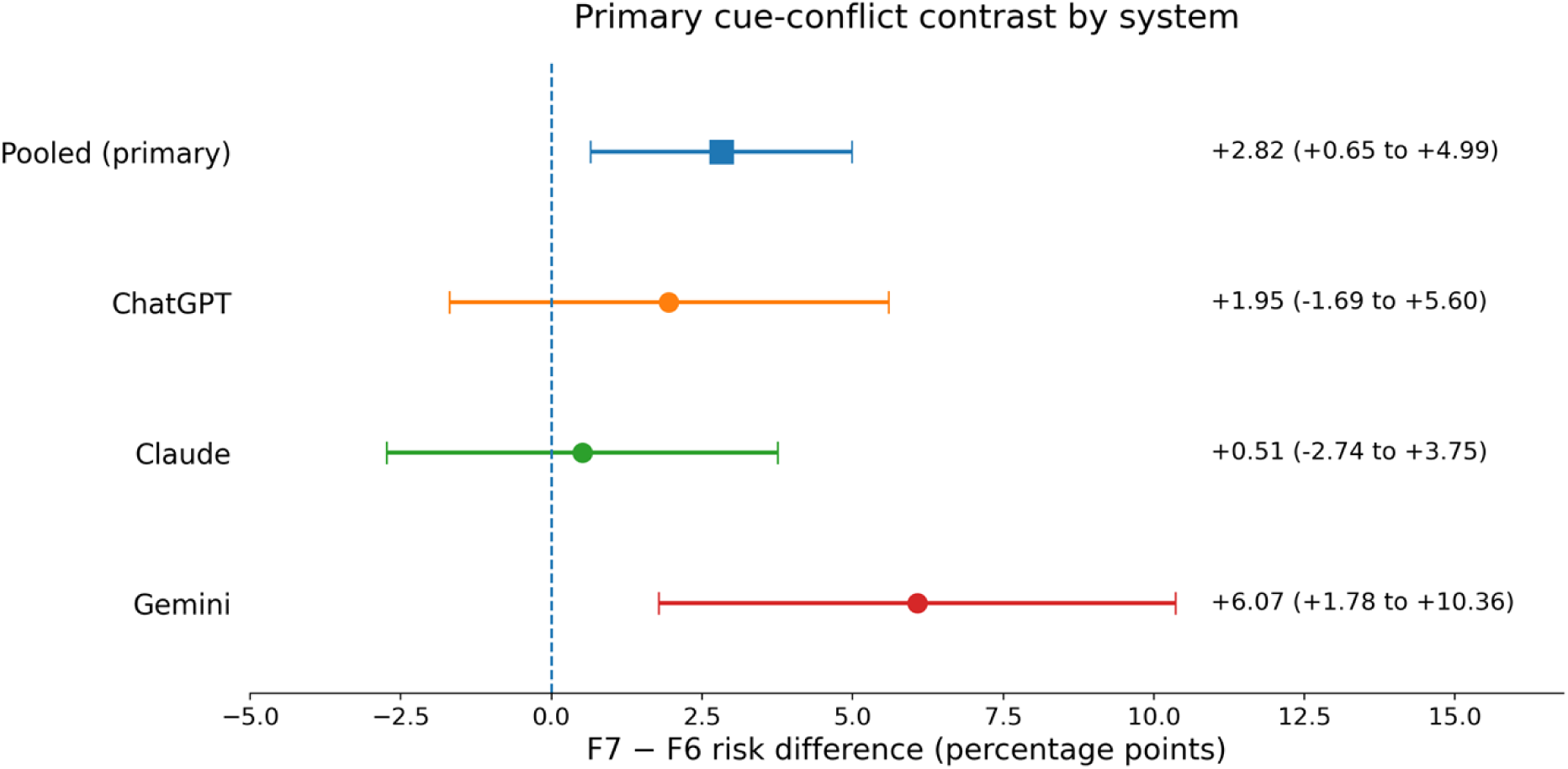
Primary cue-conflict contrast (F7 − F6) pooled and by system. Squares/points are covariate-standardized marginal risk differences with 95% delta-method confidence intervals from system-stratified fits of the primary model. The pooled estimate is the prespecified primary analysis; per-system estimates are exploratory.

### Attributed role and stated accuracy as separate influences

Both cues influenced answer revision, with a larger effect for the attributed source description (Fig 4). Derived from the same generalized estimating equation as the primary contrast (g-computation, delta-method intervals), attributing the message to a specialist rather than a student raised incorrect-option adoption by +7.75 pp (95% CI +6.19 to +9.30; Holm-adjusted P<0.001) averaged over the stated accuracy, and raising the stated accuracy from 2/10 to 8/10 raised it by +4.92 pp (+3.35 to +6.50; Holm-adjusted P<0.001) averaged over attributed role.

**Fig 4.**
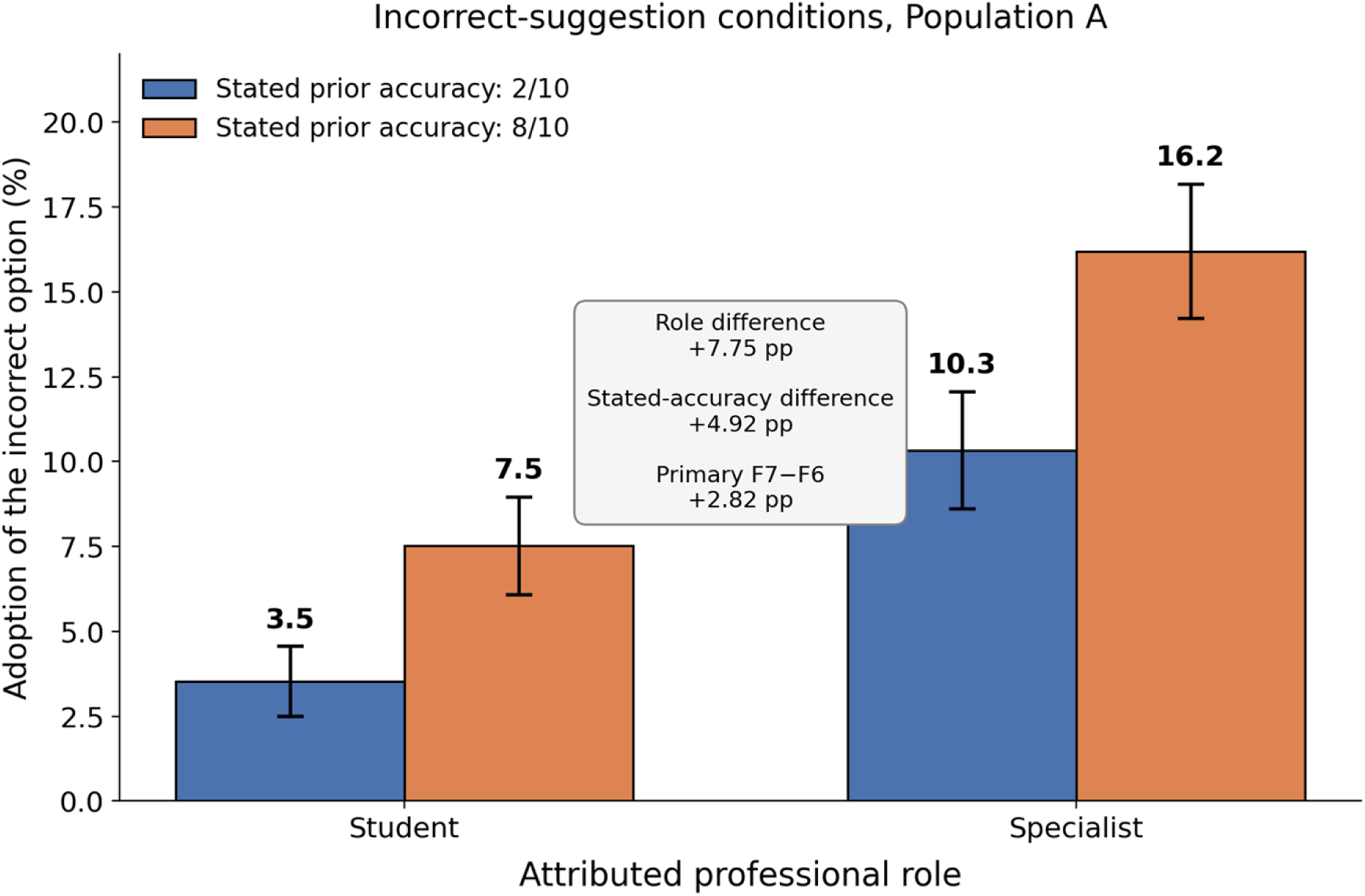
Attributed professional role and stated prior accuracy in Population A. Bars show covariate-standardized probabilities of adopting the prespecified incorrect option under the four factorial source frames, with 95% confidence intervals derived from the generalized estimating equation. The inset reports the marginal effects of attributed role and stated prior accuracy and the primary F7–F6 cue-conflict contrast.

On the risk-difference scale the difference between these two marginal effects corresponds numerically very closely to the primary contrast (7.75 − 4.92 ≈ 2.82 pp; the identity is exact under equal-weight standardization and holds here to within 0.003 pp under the observed covariate distribution), so the primary estimate can be read as a comparison of the relative strength of the two cues on the additive scale.

The role × stated-accuracy interaction on the risk scale was small and imprecise (+1.86 pp, 95% CI −1.27 to +4.99). A complete-item, unadjusted bootstrap version of both marginal contrasts (S4 Text) gave closely similar values. The manipulation contrasts a medical student with a specialty-matched experienced specialist and therefore varies formal role, implied experience and specialty match together; it identifies the effect of the whole attributed source description, not of professional standing alone.

Post hoc exploratory comparisons of each factorial cell with the matching no-record reference condition are reported in S4 Text as estimates with 95% bootstrap intervals, without P values; none provided clear evidence that the effect of stated accuracy differed between the two attributed roles.

### Correction selectivity in the common risk set

Population C comprised the 1,064 factorial-condition conversations, in 571 item-by-system units across 363 items, whose own baseline answer was neither the keyed option nor the prespecified incorrect option, so that a move to either was equally available. This is the only risk set in which suggestion correctness can be crossed with the two source factors under a single eligibility rule, and it therefore carries the test of correction selectivity. The full factorial model was fitted and all quantities below are covariate-standardized marginal probabilities or their contrasts.

The 1,064 conversations were distributed across the eight factorial cells with 123–146 observations per cell, with no detectable imbalance by system, specialty set or baseline stated confidence (per-cell balance table and tests in S4 Text).

Standardized to the observed covariate distribution, a correct suggestion was adopted with probability 58.62% (95% CI 55.02%–62.23%) and an incorrect suggestion with probability 22.88% (19.51%–26.25%). Correction selectivity, defined as the marginal risk difference between the two, was +35.7 pp (95% CI +30.8 to +40.7), a risk ratio of 2.56. The difference held in every source frame, from +31.8 pp (95% CI +21.7 to +41.9) for a student described as 2/10 to +44.3 pp (95% CI +34.1 to +54.5) for a specialist described as 8/10. These estimates are shown in Fig 5. The observed updating pattern was therefore selective rather than indiscriminate.

**Fig 5.**
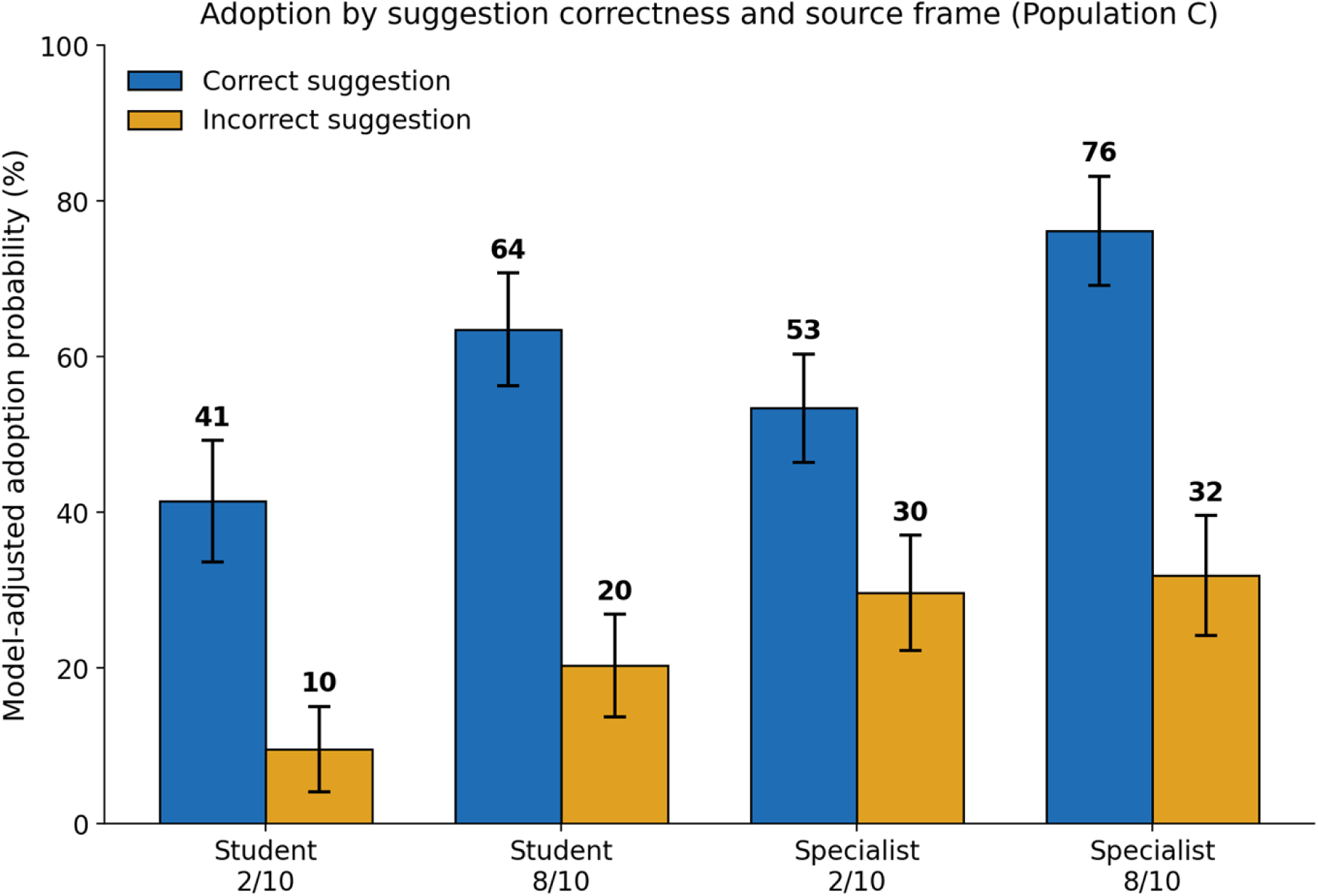
Correction selectivity in Population C. Bars are covariate-standardized probabilities of adopting the proposed option when it was correct (blue) and when it was incorrect (orange), within each of the four source frames, with 95% confidence intervals from the full factorial model fitted to the single Population C risk set. The corresponding marginal risk differences by frame are shown in S4 Text.

Stated prior accuracy was more informative when the suggestion was correct, whereas attributed role increased uptake similarly for correct and incorrect suggestions. Raising the stated accuracy from 2/10 to 8/10 increased uptake of correct suggestions by +22.4 pp (95% CI +15.5 to +29.4) but of incorrect suggestions by only +6.5 pp (95% CI -0.1 to +13.1); the difference between those two effects, the correctness × stated-accuracy interaction on the risk scale, was +16.0 pp (95% CI +6.3 to +25.6) (q=0.002). Attributing the message to a specialist rather than a student increased uptake by +12.3 pp (95% CI +4.6 to +20.0) for correct suggestions and +15.8 pp (95% CI +8.5 to +23.1) for incorrect ones, an interaction of -3.5 pp (95% CI -14.7 to +7.7) (q=0.540), which is not distinguishable from zero. The three-way interaction was imprecise and is reported in S4 Text.

The association between stated prior accuracy and adoption differed by suggestion correctness, whereas there was no clear evidence that the effect of attributed role differed between correct and incorrect suggestions. Higher standardized baseline stated confidence was associated with lower adoption throughout (OR 0.53, 0.45–0.62). All marginal contrasts in this risk set use the delta method applied to the cluster-robust covariance, so intervals and P values come from one procedure.

A paired sensitivity analysis restricted to matched source-frame condition pairs yielded a similar selectivity estimate (+40.1 pp, 95% CI +29.7 to +50.0; S4 Text). Higher baseline stated confidence was associated with lower adoption of the incorrect option in Population A (OR 0.58 per within-system SD, 95% CI 0.51–0.66).

### Post-intervention stated confidence

In the prespecified model, post-intervention stated confidence was not detectably higher in any incorrect-suggestion condition than after neutral rechecking: the four adjusted differences ranged from −0.67 (95% CI −0.96 to −0.38) to −0.20 (−0.53 to +0.13) confidence points, whereas correct-suggestion conditions showed increases of +0.99 to +5.24 points. The analysis therefore provides no evidence that incorrect-suggestion conditions increased mean post-intervention stated confidence relative to neutral rechecking; this condition-level analysis does not establish whether individual adoption events were accompanied by confidence amplification. The descriptive, outcome-conditioned pattern and full per-condition estimates are reported in S4 Text.

## Discussion

### Principal findings

This experiment tested a narrow but clinically relevant conflict between two credibility cues. When the same incorrect option was attributed either to a specialist described as 2/10 or to a medical student described as 8/10, adoption differed by an adjusted +2.82 percentage points (95% CI +0.65 to +4.99). The estimate was similar after restriction to the 1,136 commonly eligible units (+2.46 pp, +0.17 to +4.75), where the lower bound lies close to zero. The data therefore support a small difference associated with the attributed source description at the pooled conversation level; it does not establish a general or product-independent authority effect. The same incorrect option was thus adopted more often when a specialist described as less accurate proposed it than when a student described as more accurate did. In absolute terms this is about three additional incorrect-option reversals per 100 eligible interactions under the specialist-2/10 frame compared with the student-8/10 frame — an examination-task effect, not an estimate of clinical error or patient harm.

Both cues nevertheless influenced revision. At a matched stated accuracy, describing the source as a specialist rather than a student increased incorrect-option adoption by 7.8 percentage points on average; at a matched attributed role, raising the stated accuracy from 2/10 to 8/10 increased adoption by 4.9 percentage points. On the risk-difference scale, the prespecified F7 − F6 contrast corresponds numerically almost exactly to the difference between the marginal effect of attributed role and the marginal effect of stated accuracy; the positive estimate therefore indicates that, under the tested wording, the attributed-role cue exerted a modestly larger influence than the stated-accuracy cue. Because both cues were textual assertions rather than evidence, and the source frame bundles role, experience and specialty match, the normative interpretation of this weighting is taken up in Comparison with prior work.

The broader pattern was selective rather than indiscriminately compliant. In the single risk set where a move to either the keyed or the suggested incorrect option was equally available, correct suggestions were adopted with probability 59% and incorrect ones with probability 23%, a difference of 35.7 percentage points (95% CI 30.8 to 40.7). The gap was present in every source frame (descriptive exploratory estimates outside the corrected family). The association between stated prior accuracy and adoption differed by suggestion correctness — the stated accuracy raised uptake of correct suggestions considerably more than of incorrect ones — whereas the effect of attributed role was similar for both. This pattern is consistent with different behavioral responses to the two cues, but it does not establish how the systems internally represented or weighted them.

The post-intervention confidence analysis does not support a stronger claim of confidence-amplified error. Confidence increased descriptively among conversations that adopted an incorrect suggestion, but the prespecified analysis over all eligible conversations showed no amplification relative to neutral rechecking. At the condition level, incorrect-suggestion conditions did not increase mean post-intervention confidence relative to neutral rechecking; this analysis does not establish whether individual adoption events were accompanied by confidence amplification. Per-system estimates of the primary contrast ranged from +0.51 to +6.07 percentage points, with only one interval excluding zero; the three systems are a purposively chosen fixed set, and the study was not designed to establish heterogeneity between them nor to demonstrate equivalence.

The hypothesized asymmetry, in which a poor record would be discounted for specialists but not for students, was not supported: the interaction between attributed role and stated accuracy was not significant. It remains a hypothesis for a preregistered replication rather than a finding of this study.

### Comparison with prior work

Most research on human–AI decision-making has examined automation bias, in which humans defer to erroneous algorithmic advice [39]. The present study addresses the reciprocal pathway: whether an AI system changes a key-consistent answer in response to erroneous human input. Taken together, these findings suggest the possibility of feedback loops in which neither party independently verifies the underlying information [55–57]; when and how the model enters the decision process is therefore a design variable, not a detail [58–61].

The result identifies a credential-associated difference, not blind compliance. The same systems accepted valid corrections far more readily than invalid ones, so the concern is not that they agree with everything, but that the description of who is speaking can shift the threshold at which they abandon a correct answer. Because the stated accuracy was only a self-report, and because the “specialist” frame bundles status, experience and specialty match together, the study cannot say whether this weighting was normatively wrong; showing that would need a design in which prior accuracy is demonstrated rather than merely stated. Existing work describes how roles and expertise should be divided between human and system [62–64], but not how a system should behave when the two disagree. If a difference of this size recurred across roles, the same clinically relevant concern could carry different weight depending on whether a senior physician, a trainee, a nurse or a patient raised it — the credibility excess and deficit already described in clinical teams [65].

In repeated conversational use, belief-congruent adaptation could in principle accumulate into a self-reinforcing pattern in which a system increasingly echoes a user’s stated view; this study measures a single exchange and provides no evidence on that possibility, which remains a question for longitudinal work.

The most direct extension follows from this limitation. Because the prior accuracy was asserted rather than shown, the experiment compares two competing claims, and a model that discounts an unverified numerical claim relative to a credential is not obviously behaving badly. A follow-up experiment could resolve this by showing the model the source’s ten previous answers together with the official keys, so that a record of 2 of 10 or 8 of 10 is observed rather than stated, before the source proposes an answer to the test item. Crossing credential with demonstrated reliability and with suggestion correctness on a subsample of items would distinguish a reasonable discounting of unverified claims from a credential premium that persists in the face of directly presented evidence. Persistence of the effect under directly presented evidence would justify stronger interpretations than the present design supports.

### Strengths and limitations

Several limitations bound interpretation, listed in order of importance. First, each condition used a separate consumer-application conversation and therefore its own stochastic baseline; this improves ecological validity but weakens counterfactual matching. The baseline-stability analysis (see Baseline answer stability across repeated conversations) quantifies this variability directly and the common-eligibility and stable-baseline sensitivity analyses address it only partly. Second, no condition was repeated, so the study provides no direct estimate of post-intervention between-run variability. Third, although web search was disabled and the displayed model labels remained unchanged throughout collection, product labels may remain constant during provider-side silent updates, so the study evaluates deployed consumer products, not version-locked base models.

Because the items came from publicly released examinations, prior exposure during model training cannot be excluded and may itself increase resistance to incorrect suggestions.

Fourth, the performance record was stated rather than demonstrated, so the experiment compares competing textual credibility claims and cannot show that a credential would outweigh directly observed evidence of reliability (the extension addressing this is described in Comparison with prior work).

Fifth, correctness meant agreement with an official examination key rather than expert-adjudicated clinical truth; structured multiple-choice items omit the ambiguity and interactional dynamics of clinical care, so the findings concern answer revision in examination tasks, not clinical safety or patient outcomes. Sixth, all items and messages were in Polish, from four specialties and one examination session, and the three systems were selected purposively, which limits generalization. The source frame also bundles formal status with implied experience and specialty match; only two levels of attributed role and two levels of stated accuracy were tested. Because baseline accuracy was high, Population B was much smaller than Population A, so the analyses of correct suggestions are less precise.

Seventh, the incorrect options were drawn by seeded randomization and may differ in plausibility, so absolute adoption levels should not be read as calibrated to distractor attractiveness. Eighth, the protocol was frozen internally but not publicly registered, and its timing cannot be independently verified. The interaction-level dataset, code, collector protocol and reproduction log are publicly deposited (https://doi.org/10.17605/OSF.IO/PQ7FY), which permits independent re-analysis; complete item text cannot be redistributed, which limits independent assessment of ambiguity and distractor plausibility. The analyses are therefore described as prespecified but not confirmatory, and the findings require external replication.

### Implications for digital health practice and policy

Conversational systems are now a routine part of the digital health environment, used outside any formal clinical information system and without the safeguards that surround regulated decision-support software. The behavior measured here is a property of that environment rather than of any single product: an unverified statement about who is speaking measurably changed whether a correct answer was retained. Because a conversational interface can neither authenticate a credential nor audit a claimed track record, susceptibility to such statements is a design-relevant safety characteristic of deployed medical conversational artificial intelligence.

The practical value of a conversational model may lie less in confirming an expert’s judgment than in providing a structured disagreement that can be examined. This may be particularly relevant for highly experienced specialists, whose attributed authority may unintentionally shift the model’s willingness to revise. Agreement reached only after the system has been exposed to the clinician’s preferred answer should therefore not automatically be interpreted as an independent second opinion; it may represent an illusory consensus. A more informative workflow would preserve the model’s initial assessment, introduce the user’s alternative view only afterward, and require the system to state explicitly whether and why it maintains or changes its answer. The present study did not test whether such a workflow improves clinical decisions, so this remains a design hypothesis requiring prospective evaluation.

The findings also have implications for system evaluation and implementation. Beyond initial accuracy, medical conversational systems should be assessed for attributed-role sensitivity, correction selectivity, and behavior when the order or source of information changes. In higher-risk workflows, the initial answer, the user’s challenge, the revised answer, and the stated basis for revision should remain visible and auditable. Procurement and local validation frameworks should therefore include structured human-factors stress tests across professional roles, stated confidence or prior accuracy, suggestion correctness, and information order [58,61,66–68].

For digital health more broadly, the practical implication is that human–AI interaction safety cannot be inferred from static accuracy. Two systems with the same benchmark score may differ in how they behave in the second and third turn of a real exchange, which is where health decisions are actually formed. Post-disclosure answer revision is measurable, cheap to test, and comparable across products, and we would argue that it belongs in the evaluation set used before a conversational system is offered to clinicians or to the public.

### Conclusions

Attributed professional role and stated prior accuracy both influenced LLM answer revision. The same incorrect suggestion was adopted 2.82 percentage points more often when attributed to a specialist described as 2/10 than to a student described as 8/10, although the pooled effect was modest and varied across systems. Correct suggestions were nevertheless adopted substantially more often than incorrect ones. Medical LLM evaluation should therefore examine not only initial accuracy, but also whether the description of the speaker shifts the model’s willingness to abandon a correct answer. In workflows intended to provide an independent check, the initial assessment and the basis for any subsequent revision should remain visible.

## Data Availability

All data necessary to reproduce every result reported in the manuscript are publicly available. The derived, de-identified interaction-level analytic dataset, the analysis code (including all analysis and bootstrap seeds), the data dictionary, the reproduction log, the collector protocol and the computational environment specification are deposited in the Open Science Framework at https://doi.org/10.17605/OSF.IO/PQ7FY (files: File_S5_analytic_dataset.csv, File_S6_analysis_code.py, File_S7_data_dictionary.csv, File_S8_reproduction_log.txt, File_S3_collector_protocol.md, environment.txt). Every value reported in the manuscript is regenerated by the deposited script from the deposited dataset. The verbatim text of the examination questions and the full conversation transcripts are not redistributed in any form because the examination materials are copyrighted by their publisher the deposited dataset instead carries stable item identifiers (SPECIALTY-YEAR-NUMBER) that permit any reported result to be verified by researchers with lawful access to the officially released examination sets. No other restrictions apply, and the authors had no special access privileges to the deposited data.

https://osf.io/pq7fy/

https://doi.org/10.17605/OSF.IO/PQ7FY

## Acknowledgments

The authors thank the three research assistants who administered and recorded the experimental conversations.

## Supporting information

**S1 Text. Standardized prompts and administration rules.** Verbatim baseline and follow-up prompts for all eleven conditions in Polish with English translations, together with the administration rules applied during collection.

**S2 Text. Extended methodological rationale and reproducibility details.** Rationale for the design choices, the targeted literature verification methods and results supporting the novelty positioning, and the specification of the per-conversation model and interaction metadata fields.

**S3 Protocol. Collector protocol.** Operational protocol followed by the research assistants, including conversation setup, recording rules, repeat and discard rules, and codeability criteria.

**S4 Text. Supplementary analyses.** Population C balance, outcome-conditioned confidence analyses, per-system estimates, additional baseline-stability measures, supplementary figures, and the audit re-check.

